# Reproducible breath metabolite changes in children with SARS-CoV-2 infection

**DOI:** 10.1101/2020.12.04.20230755

**Authors:** Amalia Z. Berna, Elikplim H. Akaho, Rebecca M. Harris, Morgan Congdon, Emilie Korn, Samuel Neher, Mirna M’Farrej, Julianne Burns, Audrey R. Odom John

## Abstract

SARS-CoV-2 infection is diagnosed through detection of specific viral nucleic acid or antigens from respiratory samples. These techniques are relatively expensive, slow, and susceptible to false-negative results. A rapid non-invasive method to detect infection would be highly advantageous. Compelling evidence from canine biosensors and studies of adults with COVID-19 suggests that infection reproducibly alters human volatile organic compounds (VOCs) profiles. To determine whether pediatric infection is associated with VOC changes, we enrolled SARS-CoV-2-infected and -uninfected children admitted to a major pediatric academic medical center. Breath samples were collected from children and analyzed through state-of-the-art GCxGC-ToFMS. Isolated features included 84 targeted VOCs. Candidate biomarkers that were correlated with infection status were subsequently validated in a second, independent cohort of children. We thus find that six volatile organic compounds are significantly and reproducibly increased in the breath of SARS-CoV-2-infected children. Three aldehydes (octanal, nonanal, and heptanal) drew special attention, as aldehydes are also elevated in the breath of adults with COVID-19. Together, these biomarkers demonstrate high accuracy for distinguishing pediatric SARS-CoV-2 infection and support the ongoing development of novel breath-based diagnostics.

## INTRODUCTION

Novel diagnostic strategies are urgently needed to control the current COVID-19 pandemic, caused by infection with the novel coronavirus SARS-CoV-2. SARS-CoV-2 infection is typically diagnosed through detection of viral specific nucleic acids or antigens from samples collected from the upper respiratory tract (e.g. saliva, oropharyngeal, nasal, or nasopharyngeal swabs). Such testing can be uncomfortable and relatively expensive, requires multiple reagents for which supplies can be limited (swabs, viral transport media, and RNA extraction kits), and relies on specialized laboratory equipment and trained personnel. Moreover, high false-negative rates have been reported.^1^ Improved access to simple, rapid diagnostic testing for SARS-CoV-2 infection would facilitate COVID-19 control efforts and support clinical care of symptomatic or exposed individuals, especially in resource-limited environments. In addition, a rapid diagnostic test suitable for large-scale screening of children for coronavirus infection could facilitate infection prevention and control in congregate living or educational settings.

Children with SARS-CoV-2 infection typically experience mild symptoms of disease and are much less likely to experience severe outcomes, such as hospitalization or death, as compared to adults.^2^ Children also exhibit a distinct immunological response to coronavirus infection.^3-4^ These clinical and immunological differences in children with SARS-CoV-2 suggest that their metabolic response to infection may also be different than that of adults. However, any novel diagnostic test for SARS-CoV-2 will require excellent performance characteristics in children, due to the importance of testing in this population. Asymptomatic or mildly symptomatic pediatric cases may transmit SARS-CoV-2 within the household or community.^5^ While adults are inconvenienced by social distancing measures to control viral transmission, the educational and social development of children may be irreparably harmed.^6^ Finally, until global control of COVID-19 is achieved, children will continue to require a disproportionately high frequency of testing, due to both the burden of clinically indistinguishable non-SARS-CoV-2 upper respiratory viral infections in childhood (up to 12 per year) and the delayed availability of vaccination for young children.^7^

Metabolic changes induced by respiratory infections may alter host odor profiles, such that infection-associated volatile organic compounds (VOCs) may be used to develop noninvasive diagnostics through sensor arrays (e.g. “breathalyzers”) or electronic noses. Promising proof-of-concept comes from studies of other respiratory infections that lead to characteristic alterations in breath metabolite profiles, including infection with *Mycobacterium tuberculosis*^8^ and *Aspergillus* spp., as well as ventilator-associated pneumonia.^9^ Viral respiratory pathogens impact host volatile production *in vitro* and *in vivo*. For example, infection with either rhinovirus^10^ or influenza in cell culture^11^ results in reproducible VOC changes. Similarly, studies in an animal model of influenza infection demonstrate an increase in breath concentrations of acetaldehyde, propanal, and n-propyl acetate.^12^ More recently, compelling evidence from canine biosensors suggests that volatile detection may be a promising approach for SARS-CoV-2 diagnosis. Trained dogs reproducibly recognize SARS-CoV-2 infection in saliva/tracheal samples^13^, urine^14^ and sweat samples.^15^ In addition, distinct breath signatures were found in adult patients with COVID-19, compared to those with unrelated respiratory and cardiac conditions.^16^ Preliminary studies using sensor arrays confirm that SARS-CoV-2 infection in adults likewise results in distinct breath volatile changes.^17^

To evaluate whether changes in breath metabolites also characterize the exhaled breath of pediatric patients with SARS-CoV-2 infection, we analyzed breath metabolite profiles from two independent cohorts of children with and without SARS-CoV-2, who were admitted to a major pediatric academic medical center (for workflow, see Supplemental Figure 2). Through targeted GCxGC-mass spectrometric analysis of 84 breath volatiles, we established candidate biomarkers of pediatric SARS-CoV-2 infection, which were validated in an independent cohort of children.

## RESULTS AND DISCUSSION

Biomarker discovery was performed from breath metabolic profiling of pediatric patients (n=26) from the Children’s Hospital of Philadelphia (CHOP), 11 of whom were positive and 15 of whom were negative for SARS-CoV-2 by nasopharyngeal (NP) RT-PCR. One SARS-CoV-2-infected subject was excluded due to poor quality of breath sampling.

Demographic and clinical characteristics in this discovery cohort are shown in Table 1. SARS-CoV-2-infected and -uninfected patients were broadly similar with respect to age, sex, and racial/ethnic characteristics. Individuals infected with SARS-CoV-2 were more likely to exhibit either fever (50% vs. 0.0%, p=0.004) or cough (40% vs. 0.0%, p=0.016), compared to uninfected subjects. Two SARS-CoV-2-positive subjects (25%) lacked symptoms of acute infection (specifically fever, sore throat, cough, or GI symptoms). Two subjects with positive nasopharyngeal testing for SARS-CoV-2 were subsequently diagnosed with multisystem inflammatory syndrome in children (MIS-C), believed to be a late complication of SARS-CoV-2 infection.

**Table 1.** Demographics. Patient demographics and clinical characteristics.

|  | Discovery cohort |  |  | Validation cohort |  |  |
| --- | --- | --- | --- | --- | --- | --- |
| Variables | SARS-CoV-2 negative<br>(n = 15) | SARS-CoV-2 positive<br>(n = 10) | p value | SARS-CoV-2 negative<br>(n = 12) | SARS-CoV-2 positive<br>(n = 12) | p value |
| **Demographic characteristics** |  |  |  |  |  |  |
| Age (years), median (IQR) | 15 (12-16) | 11 (8.2-17) | 0.15 | 15 (14-16) | 12.5 (9.25-15.75) | 0.022 |
| Female, n (%) | 9 (60) | 6 (60) | >0.99<sup>1</sup> | 9 (75) | 6 (50) | >0.40<sup>1</sup> |
| Black or African-American, n (%) | 6 (40) | 5 (50) | 0.69<sup>1</sup> | 5 (42) | 4(33) | >0.99<sup>1</sup> |
| BMI/Age percentile, median (IQR) | 86 (62.5-98)<sup>2</sup> | 61.5 (45-84) | 0.11 | 26 (21-31) | 19 (17-26)<sup>3</sup> | 0.09 |
| **Reported Symptoms, n (%)** |  |  |  |  |  |  |
| Fever (>38.0°C) | 0 (0) | 5 (50) | 0.004<sup>1</sup> | 0(0) | 2(17) | 0.47<sup>1</sup> |
| Cough (new onset or worsening of chronic cough) | 0 (0) | 4 (40) | 0.016<sup>1</sup> | 0(0) | 0(0) | >0.99<sup>1</sup> |
| Sore throat | 0 (0) | 1 (10) | 0.40<sup>1</sup> | 0 (0) | 1(8) | >0.99<sup>1</sup> |
| Headache | 0 (0) | 1 (10) | 0.40<sup>1</sup> | 0 (0) | 1(8) | >0.99<sup>1</sup> |
| **Laboratory** |  |  |  |  |  |  |
| Cycle threshold values (SARS-CoV-2 RT-PCR), median (IQR) | >40 (negative) | 36.74 (31.78-37.63) | --- | >40 (negative) | 28.83 (22.15-32.92)<sup>4</sup> | --- |

Data represent median value (interquartile range) or number of patients (%).
<sup>1</sup> Fisher's exact test used for contingency table analysis. <sup>2</sup> Data unavailable for two patients. <sup>3</sup> Data unavailable for one patient.
<sup>4</sup> Data unavailable for four patients.

For each patient, breath volatiles were captured onto sorbent material and subsequently released by thermal desorption for analysis by two-dimensional gas chromatography and time-of-flight mass spectrometry (GCxGC ToF-MS). Isoprene is one of the most common and abundant human breath VOCs. To establish the quality of breath VOC collection, the abundance of isoprene was compared to the abundance of isoprene in ambient air, which had been collected in the same room and at the same time as breath collection. For each subject, we find that the abundance of isoprene was markedly higher than ambient levels, confirming successful breath VOC collection (Supplementary Figure 3).

For our targeted metabolite analysis, we selected 84 VOCs that have previously been identified as either common human odorants^18^, associated with host response to viral infection, or were found to be elevated in the breath of adults with COVID-19.^10-12, 16, 19-20^ Volcano plots (Figure 1a) were used to visualize breath metabolic features that distinguished SARS-CoV-2-infected from - uninfected individuals, using p<0.05 as a threshold for statistical significance. Six candidate breath biomarkers were significantly elevated in the breath of children with SARS-CoV-2 infection: three aldehydes [octanal, nonanal, and heptanal (Figure 2)], as well as decane, tridecane, and 2-penthyl furan (Figure 1b and Supplementary Figure 4). All compound identities were confirmed by comparison to pure commercial standards. Analytical characteristics of candidate breath biomarkers can be found in Supplementary Table 1.

**Figure 1.**
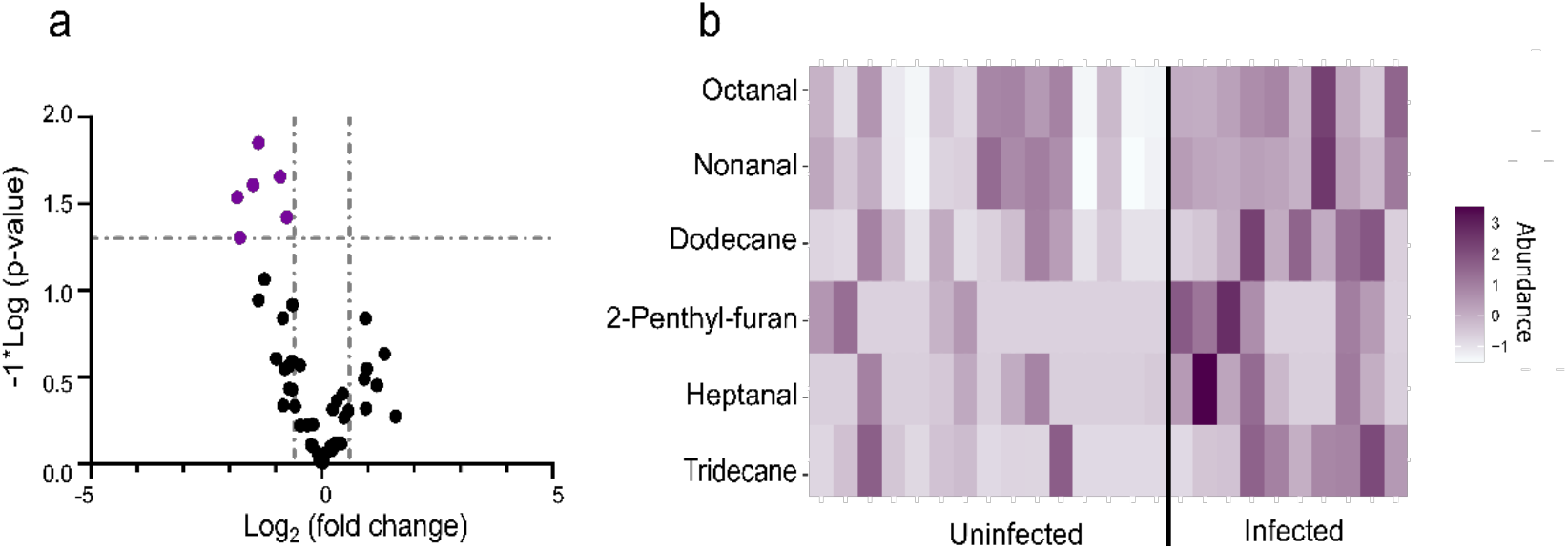
Candidate breath biomarkers of pediatric SARS-CoV-2 infection. (a) Volcano plot of breath metabolites. Fold change = mean abundance SARS-CoV-2-infected/mean abundance uninfected. Purple, breath metabolites significantly different (p<0.05) in SARS-CoV-2-infected subjects compared to uninfected. Metabolite identities are shown in heat map. (b) Heat map visualizing abundance of breath biomarkers (presented as z-scores) in a discovery cohort of uninfected- and SARS-CoV-2-infected pediatric subjects.

**Figure 2.**
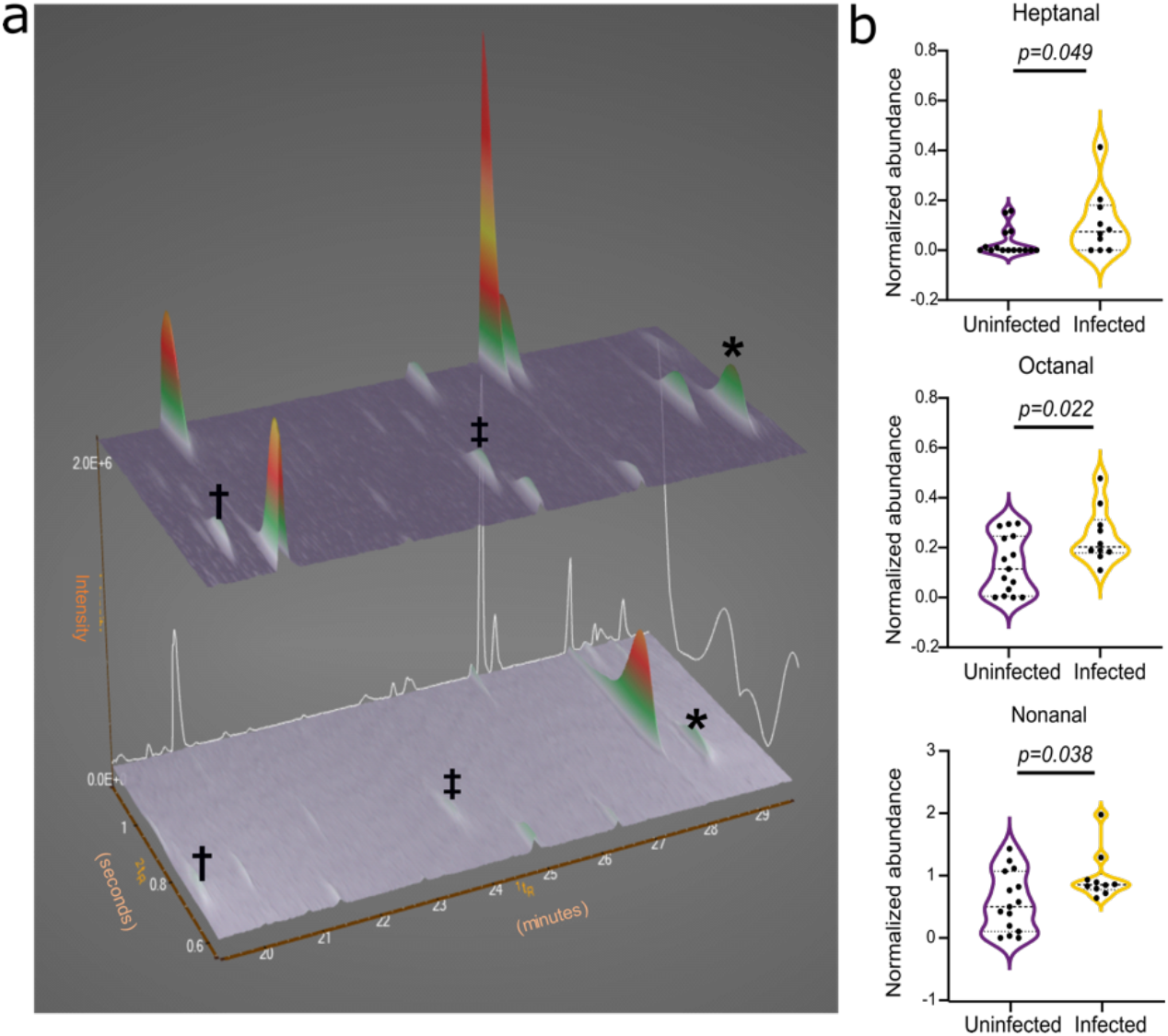
Pediatric SARS-CoV-2 infection is associated with an increased abundance of breath aldehydes. (a) Three-dimensional GCxGC ToF-MS surface plots of two representative pediatric breath samples (top, SARS-CoV-2-infected; bottom, SARS-CoV-2-uninfected), demonstrating increased abundance of characteristic medium-chain aldehydes (†, heptanal; ‡, octanal; *, nonanal) associated with SARS-CoV-2 infection. 1tR, retention time in minutes; 2tR, retention time in seconds. (b) Scatter plot of breath abundance of candidate SARS-CoV-2 aldehyde biomarkers in uninfected and infected children. Octanal and heptanal were also previously found to be increased in abundance in the breath of adults with COVID-19^16^. Median and quartiles are shown. P-values (t-tests) are shown for each comparison.

Heat map visualization indicates an increase in the abundance of candidate volatile biomarkers in the breath of children with SARS-CoV-2 infection (Figure 1b), suggesting that SARS-CoV-2 infection alters the overall profile of breath VOCs. Because elevated temperature alone can alter metabolic profiles, we evaluated whether any candidate biomarkers correlated with fever. We find that fever was not associated with significant changes in abundance of any SARS-CoV-2 biomarker (Supplementary Figure 5).

To establish the reproducibility of these candidate biomarkers, independent subjects were enrolled in a validation cohort of children with and without SARS-CoV-2 infection (n=24). Patients enrolled had similar demographic and clinical characteristics as the discovery cohort, and infected and uninfected children were broadly similar (Table 1). We found that all candidate SARS-CoV-2 biomarkers were increased in abundance in infected, compared to uninfected, children in this validation cohort (Supplementary Figure 6). To visualize the discriminatory power of these biomarkers, principal components analysis (PCA) was performed (Figure 3). This technique indicates substantial differences in breath volatile composition in the validation cohort between SARS-CoV-2-infected children compared to children without infection. Using the sum of the abundances of these 6 biomarkers (“cumulative abundance”) as a diagnostic strategy, we evaluated its diagnostic characteristics. A receiver operating characteristic (ROC) curve yielded an area under the curve (AUC) of 0.92, providing a sensitivity of 91% and specificity of 75% (Figures 4a, b and c). Overall, our results suggest that SARS-CoV-2 infection leads to characteristic and reproducible changes in breath volatiles in children, and that as few as 6 volatiles may be used to diagnose SARS-CoV-2 with high accuracy.

**Figure 3.**
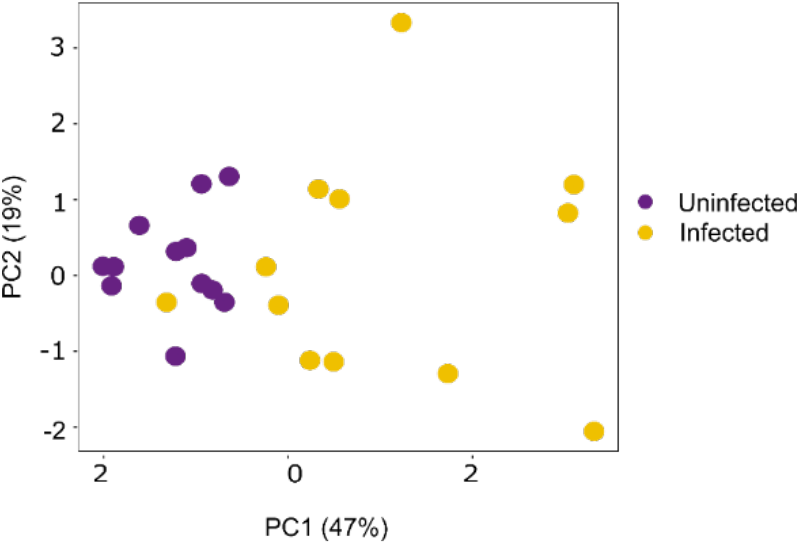
Discriminatory power of candidate SARS-CoV-2 breath biomarkers in an independent cohort of children. Principal component analysis was performed with six candidate biomarkers of SARS-CoV-2 using breath samples from an independent validation cohort of children with and without SARS-CoV-2 infection.

**Figure 4.**
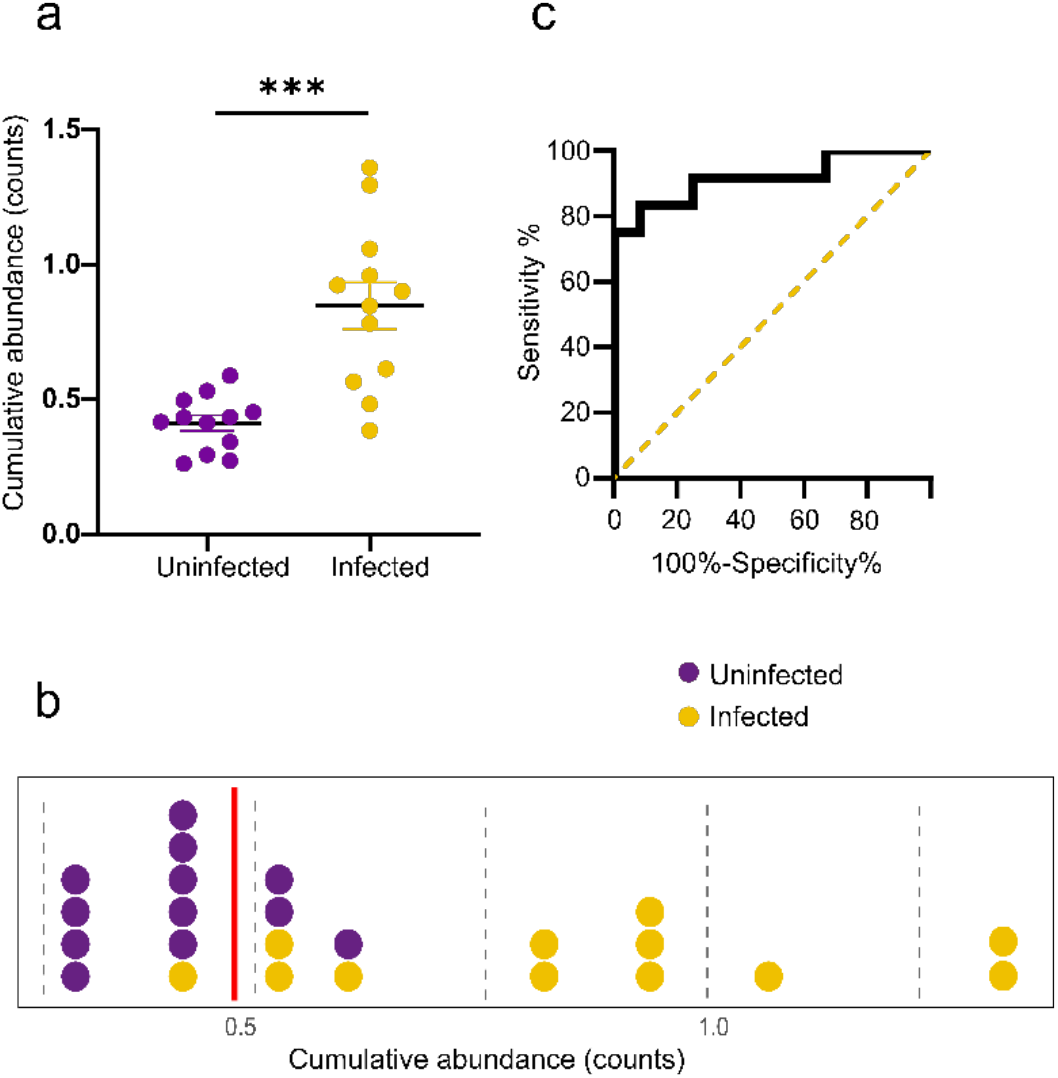
Performance characteristics of a cumulative abundance metric for SARS-CoV-2 diagnosis. (a) The cumulative abundance (normalized to internal standard) of six candidate biomarkers readily distinguishes breath profiles from an independent validation cohort of children with and without SARS-CoV-2 infection (t-test, p=0.001). The cumulative abundance is the sum of abundances of the six candidate biomarkers. (b) Distribution of cumulative abundance of biomarkers from SARS-CoV-2-infected and uninfected children. Red line, threshold of discrimination between infected and uninfected. (c) Receiver operator characteristics (ROC) curve for the cumulative abundance of 6 biomarkers. Dotted line indicates expected results if predictive power is no better than random chance. Using threshold, this cumulative abundance metric yields 91% sensitivity and 75% specificity.

Compared to adults, children have a distinct immune response and are less likely to become seriously ill with SARS-CoV-2 infection. For this reason, biomarkers enriched in the breath of adults with symptomatic COVID-19 may be distinct from those in children. We investigated whether breath volatiles that were previously found to be associated with adult COVID-19 were also present in our pediatric samples.^16^ We find that two medium-chain aldehydes that are elevated in the breath of adults with COVID-19, octanal and heptanal, are also elevated in the breath of children with SARS-CoV-2 infection (Figure 2b). Interestingly, pediatric SARS-CoV-2 infection was also associated with increased levels of a third aldehyde, nonanal. In contrast, two chemically distinct breath biomarkers of SARS-CoV-2 infection in adults, acetone and 2-butanone, are not significantly altered in SARS-CoV-2-infected children, compared to uninfected controls (Figure 5 and supplementary Figure 7).

**Figure 5.**
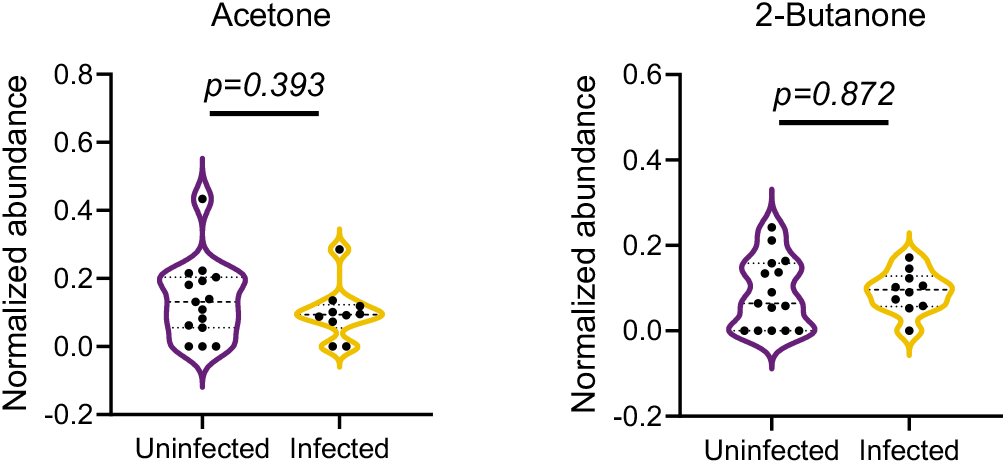
Ketones associated with adult COVID-19 are not enriched in the breath of children with SARS-CoV-2 infection. Acetone and 2-butanone were previously reported to be enriched in the breath of adults with COVID-19 ^16^. No significant differences were found between SARS-CoV-2 infected and uninfected pediatric samples in the pilot data (shown). Similar results were observed in an independent validation cohort (Supplementary Figure 7). Median and quartiles are shown. P-values (t-tests) are shown for each comparison.

Frequent, rapid testing has been proposed as an important public health strategy for control of the current COVID-19 pandemic. An easy-to-use SARS-CoV-2 “breathalyzer” based on electronic nose technologies or sensor arrays would have a turnaround time of minutes and would not strain supply chains of specialized disposable supplies. Because of ongoing advances in portable, low-cost, field-stable sensor array platforms that may be harnessed for a VOC-based diagnostic, there is enthusiastic industry support that may rapidly translate volatile biomarkers into physical devices for point-of-care testing or screening.

Strong evidence indicates that SARS-CoV-2 infection in adults leads to a unique human odorant profile. Canine biosensors (sniffer dogs) accurately recognize SARS-CoV-2 infection in biological samples and have begun to be deployed for human screening in real-world settings such as airports and sports arenas.^13, 15^ Previous studies by Ruszkiewicz et al ^16^ also report breath biomarkers associated with COVID-19 in adults presenting for emergency room evaluation with acute respiratory symptoms. They find that breath biomarkers distinguished patients with COVID-19 from those with other respiratory conditions with high (>80%) accuracy. ^16^ Metabolites, including breath volatile organic compounds, that are associated with viral infection are most likely to arise from changes in host metabolism, raising the possibility that populations that differ in their clinical response to infection might also have divergent metabolic profiles. Since pediatric SARS-CoV-2 infection is generally mild and less likely to result in serious respiratory symptoms, we expected that the breath metabolic biomarkers of children infected with SARS-CoV-2 might differ from those found in adults. In this work, we provide compelling support that SARS-CoV-2 infection leads to characteristic volatile organic compound changes in the breath of children, as it does in adults. However, we find that the breath volatile signature of pediatric SARS-CoV-2 is distinct. For example, pediatric SARS-CoV-2 infection does not lead to changes in some specific breath biomarkers, such as acetone and 2-butanone, that are highly characteristic of COVID-19 in adults.

This work provides important validation that SARS-CoV-2 infection leads to changes in breath aldehyde concentrations in both children and adults. The prior study by Ruszkiewicz et al ^16^ found that increased levels of two aldehydes, octanal and heptanal, were present in the breath of adults with SARS-CoV-2 infection, compared to those with other acute respiratory illnesses (such as COPD and pneumonia). We find that octanal, heptanal, and the structurally similar aldehyde nonanal, are all significantly elevated in the breath of children with SARS-CoV-2 infection, suggesting a common biological origin for this class of SARS-CoV-2-associated breath volatiles. Altogether our studies highlight the need to include pediatric samples in early discovery efforts to develop new and much-needed diagnostics for SARS-CoV-2, in order to identify biomarkers that are shared across all relevant populations.

SARS-CoV-2-associated breath biomarkers that are shared by adults and children may reflect a more generalized host response to viral infection. For this reason, an important limitation of contemporary biomarker discovery efforts for COVID-19, including our study, is the relatively reduced incidence of non-SARS-CoV-2 circulating respiratory viruses due to interventions such as school closures, masking, and social distancing. Other viral infections may induce host aldehyde production, an observation that has been attributed to infection-associated cellular oxidative stress that leads to oxidation of unsaturated fatty acids and accumulation of by-products. ^21-22,23^ For example, in a study of influenza A-induced breath volatiles, the breath abundance of acetaldehyde, propanal, and n-propyl acetate increased during acute infection and decreased as infection resolved.^12^ More recently, *in vitro* studies have demonstrated that infection with the seasonal coronavirus HCoV-229E leads to a significant increase in the levels of cellular fatty acids.^24^ Viral infections outside the respiratory tract may trigger similar metabolic changes, as fecal volatiles from children with rotavirus gastroenteritis were enriched in 2,3-butanedione, octanal, nonanal, and 2-heptenal, compared to samples from children without rotavirus infection.^25^ Understanding the biological origin of viral-induced aldehydes will be crucial to establishing the specificity and clinical utility of SARS-CoV-2-associated biomarkers.

## CONCLUSIONS

Increasing evidence suggests that SARS-CoV-2 infection in adults is associated with specific changes in volatile production. This study provides additional support that the breath abundance of six volatile organic compounds (including aldehydes) are altered in children with SARS-CoV2 infection. Importantly, most SARS-CoV-2-infected subjects enrolled demonstrated mild symptoms of infection and were only incidentally found to be infected due to routine pre-admission screening at our institution. Given the cost, discomfort, and false-negative results associated with RT-PCR- or antigen-based tests, breathalyzer testing for SARS-CoV-2 may provide an inexpensive, non-invasive, rapid, and highly sensitive alternative for population-based frequent screening of children.

## METHODS

### Study Approval and Enrollment

Prior to enrollment, the study was approved by the Children’s Hospital of Philadelphia (CHOP) Human Research Ethics Committee (IRB 20-017503) and by the CHOP Institutional Biosafety Committee (IBC 19-000145) for handling of human samples potentially containing SARS-CoV-2. Breath samples were collected from children (4-18 years of age) hospitalized in the Special Isolation Unit at CHOP, who had been diagnosed as SARS-CoV-2 positive by nasopharyngeal swab RT-PCR on admission (n=11). Samples from uninfected individuals were obtained from nasopharyngeal RT-PCR-negative subjects, enrolled from the Emergency Department Extended Care Unit of the Children’s Hospital of Philadelphia (n=15). Samples were collected between June and August 2020. The viral load of patient nasopharyngeal swab samples was estimated by cycle threshold value (Ct-value) of the N gene, with lower Ct-values indicating a higher viral load. Samples were considered positive if the Ct-value was ≤40, and Ct values of positive test results were obtained (Table 1). For validation studies, we collected an additional set of samples from SARS-CoV-2-infected children (n=12) and from SARS-CoV-2-uninfected individuals (n=12). Samples were collected as above from CHOP between October 2020 and March 2021. The sample size for this cohort was based on a calculated effect size of 1.5 between SARS-CoV-2-infected and -uninfected samples for the 6 biomarkers, which predicted that 12 samples in each group would yield a power (1-β error prob) of 0.97 (p < 0.05).

Exclusion criteria for control subjects included current rhinorrhea, cough, or diarrhea, in order to exclude individuals that may have false negative SARS-CoV-2 testing. In addition, subjects were excluded if they required oxygen supplementation within 3 hours of breath sample collection. Samples were not screened for common circulating non-SARS-CoV-2 human coronaviruses or other viral respiratory pathogens.

### Breath sample collection

Breath collection was performed as previously described^26-27^. In brief, SARS-CoV-2-infected and -uninfected subjects exhaled through a disposable cardboard mouthpiece connected to a chamber. The chamber was then attached using tubing to a 3-L SamplePro FlexFilm sample bag (SKC Inc, Pennsylvania) (Supplementary Figure 1). The volunteers were asked to take a few deep breaths, place the cardboard tube between the lips, and exhale completely. Neither a nose clip nor VOC filter were used. Breath from the bags was transferred to a sorbent tube as previously described^26, 28^. Briefly, 1 L of the breath sample was transferred to sorbent tube at 200 mL min^-1^ using an electric pump, so that all tubes had consistently the same volume. Three-bed Universal sorbent tubes containing Tenax, Carbograph, and Carboxen were used (Markes International Limited, UK). For each participant, ambient air samples and breath samples were collected from the same room. Samples were stored at 4°C until the time of analysis (within 2 weeks of collection). Samples were analyzed using thermal desorption and GCxGC BenchTOF-MS (SepSolve Analytical, UK). Analytical parameters are described in Supplementary Material.

## Data Availability

All data referred to in the manuscript is included in the figures of the manuscript. Supplemental material is also included in the link

## AUTHOR CONTRIBUTIONS

**Amalia Z. Berna**. Project coordinator, assisted with ethical approval, recruitment, sample collection, instrumental analysis, data interpretation and writing the manuscript.

**Elikplim H. Akaho**. Assisted with ethical approval, recruitment, sample collection and revision of the manuscript

**Rebecca M. Harris**. Assisted with data collection and revision of manuscript

**Morgan Congdon**. Sample collection, revision of manuscript.

**Emilie Korn**. Sample collection, revision of manuscript

**Samuel Neher**. Sample collection, revision of manuscript

**Mirna M’Farrej**. Assisted with subject recruitment, revision of manuscript

**Julianne Burns**. Sample collection, clinical coordination with the Special Isolation Unit, revision of manuscript and ethical approval

**Audrey R. Odom John**. Study conception, study design, assisted with ethical approval, data interpretation, and manuscript writing.

## ACKNOWLEDGMENTS

This work was supported by the National Institutes of Health [grant numbers: R61-DH105594, R01-AI103280, R21-AI123808, and R21-AI130584], and AOJ is an Investigator in the Pathogenesis of Infectious Diseases (PATH) of the Burroughs Wellcome Fund. We express our gratitude to the children and families of the Children’s Hospital of Philadelphia for all their support and participation. We acknowledge the unique efforts of the front-line clinical staff to facilitate breath collection.

