## Supplemental methods and supplemental data for "Reproducible breath metabolite changes in children with SARS-CoV-2 infection"

### Thermal desorption and GCxGC parameters

Prior to analysis, sorbent tubes were brought to room temperature and loaded into autosampler (Utra-xr, Markes International, UK). A gaseous standard mixture (1.01 ppm Bromochloromethane, 1.04 ppm 1,4-Difluorobenzene, 1.04 ppm Chlorobenzene-D5, 0.96 ppm 4-bromofluorobenzene) was immediately added to each tube, followed by a purge pre-desorption step consisting of 10 min with He at 50 mL*min1, to remove water content in breath samples. Tubes were thermally desorbed for 10 min at 270°C (Unity-xr, Markes International, UK) and transferred to a "Universal" cold trap which matched the sorbent of the sample tube, held at 10°C and subsequently heated to 300°C, to minimize band broadening. The split flow after the cold trap was 15 mL*min-1.

Analysis by two-dimensional gas chromatography was conducted using an Agilent 7890B GC system, fitted with a flow modulator and a three-way splitter plate coupled to a flame ionization detector and a time-of-flight mass spectrometer with electron ionization (SepSolve, UK). Chromatographic analysis was performed using a Stabilwax (30 m × 250 μm ID × 0.25 μm df) as the first dimension (1D)-GC column and a Rtx-200 MS (5 m × 250 μm ID × 0.1 μm df) as second dimension (2D)-GC column, both purchased from Restek (Bellefonte, PA, US). The following GC oven temperature program was used: initial temperature 40°C and held for 1 min, ramped to 260°C at 3°C*min-1. The final temperature of 260°C was held for 1 min. The total run time for the analysis was 75 min. Helium carrier gas was flowed at a rate of 1.2 mL*min-1. The flow modulator (Insight, SepSolve Analytical, UK) had a loop with dimensions 0.53 mm i.d. x 110 mm length (loop volume: 25 uL), and the modulation time was 2 s total.

**TOF-MS Conditions**

The GCxGC was interfaced with a BenchTOF-select time-of-flight mass spectrometer (SepSolve Analytical, UK). The acquisition speed was 50 Hz and mass range was 30-400 m/z. The ion source and transfer line were set at 250 °C and 270 °C respectively and filament voltage at 1.6 V. Electron ionization energy was 70 eV. ChromSpace (SepSolve Analytical, UK) was used to synchronize and control the INSIGHT modulator, thermal desorption, GC, and TOF.

**Chemical standards and solutions**

Nonanal, octanal, heptanal, tridecane, and 2-pentylfuran and isoprene were purchased from Sigma-Aldrich (St. Louis, MO, US). Dodecane was purchased from Merck (Darmstadt, Germany). To spike the compound of interest into a sorbent tube, a 10 ppm solution was prepared in HPLC grade methanol. Using a solution loading rig (Markes International Limited, UK), 1 μL of the solution was spiked into a sorbent tube. The sorbent tube was flushed for 3 min with nitrogen at a flow of 100 mL.min-1. All the stock solutions were stored in glass vials and kept at 4 °C. Sorbent tubes containing standards were analyzed by GCxGC BenchTOF-MS following the same protocols as described below for breath samples.

**Quality control**

Breath concentration of the canonical human volatile isoprene was performed to quality control for correct breath sampling, as a small or missing isoprene peak indicates an error in the sample collection and/or analysis, resulting in data being excluded. To check for changes in instrument sensitivity over time, a mixture of external standards was analyzed with the GCxGC BenchTOF-MS alongside the breath samples as described previously(1). Briefly, we analyzed an external standard before running each batch of breath samples. The standard used was EPA 8240B Calibration Mix (2-butanone, isobutanol, 4-methyl-2-pentanone and 2-hexanone). One mL 2000 μg.mL-1 vial standards were purchased from Sigma-Aldrich. To spike the mixture into a sorbent tube, a 100 μg.mL-1 solution was prepared in HPLC grade methanol. Using a solution loading rig (Markes International Limited, UK), 1μL of the solution was spiked into a sorbent tube. The sorbent tube was flushed for 3 min with nitrogen at a flow of 100 mL.min-1 and analyzed by GCxGC BenchTOF-MS.

**Data processing and statistical analyses**

Data was acquired and processed using ChromSpace (SepSolve Analytical, UK). All statistical analyses were performed using RStudio v1.3.1073 (PBC, Boston, MA) and GraphPad Prism V.8.4.3 (GraphPad Software, San Diego, CA). The workflow for data processing and statistical analysis is shown in Supplementary Figure 2. Background from the raw BenchTOF data file was removed using ChromSpace, and the Dynamic Background Compensate (DBC) of 0.2s peak width and noise factor 6.9 for typical GCxGC data was applied. DBC files were then integrated using the following parameters: peak detection deconvolution algorithm with a minimum ion count of 2000, absolute minimum peak area was set at 15,000 counts, absolute minimum peak high was set at 10,000 counts, and no relative threshold was set for either mass height or absolute area. Compounds were given annotations using the NIST v.17 reference library. Deconvoluted peaks were exported into .xls format file. The data were then processed using RStudio to generate integrated signal for every isolated feature. Isolated features included 84 targeted volatiles, as described below.

We targeted volatiles that have been previously associated with respiratory viruses from cell culture, from analysis of *in vitro* airway cells infected with human rhinovirus, *in vivo* breath profile in swine during Influenza A infection, previously associated breath markers of COVID-19 in adults, known human body volatiles (2-8), and authors' own unpublished breath VOC library (Supplementary Table 2). The data included three internal standards (Supplementary Table 2). Chromatographic data was first normalized using internal standard (1,4-difluorobenzene), and a volatile was retained if it was present in more than 50% of the samples in either group (i.e. infected or uninfected). In total, 50 VOCs were retained and used for further statistical analysis. Unpaired Student's t-test was used to identify metabolites that were significantly different between control groups and SARS-CoV-2 groups, with a p-value of 0.05 established as the threshold for statistical significance. Of note, multiple comparison corrections of metabolomic data can increase type II errors, because metabolites are typically highly correlated and not independent features(9, 10).

Six promising breath biomarkers of SARS-CoV-2 infection were identified through volcano plot of p-value versus fold change (mean abundance SARS-CoV-2-infected/mean abundance uninfected). Principal component analysis (PCA) was used to visualize the variance between samples in the validation set given our selection of important biomarkers. The discriminative power was assessed by ROC curve.

**Supplementary Table 1.** VOC biomarkers selected as best discriminants between SARS-Cov-2 positive and control (SARS-Cov-2 negative) patients, together with analytical characteristics of each compound.

| Compound Name | Formula | Structure | NIST match | $Rt_1$ | $Rt_2$ |
|---|---|---|---|---|---|
| **Octanal** | $C_8H_{16}O$ | 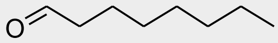 | 804 | 24.3 | 0.73 |
| **Nonanal** | $C_9H_{18}O$ | 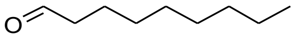 | 839 | 28.8 | 0.73 |
| **Dodecane** | $C_{12}H_{24}$ | 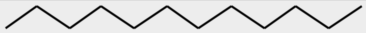 | 846 | 19.8 | 1.18 |
| **Furan, 2-penthyl** | $C_9H_{14}O$ | 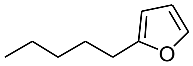 | 674 | 21.5 | 0.49 |
| **Heptanal** | $C_7H_{14}O$ | 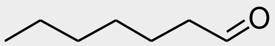 | 762 | 19.7 | 0.7 |
| **Tridecane** | $C_{13}H_{28}$ | 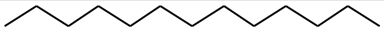 | 801 | 24.7 | 1.11 |

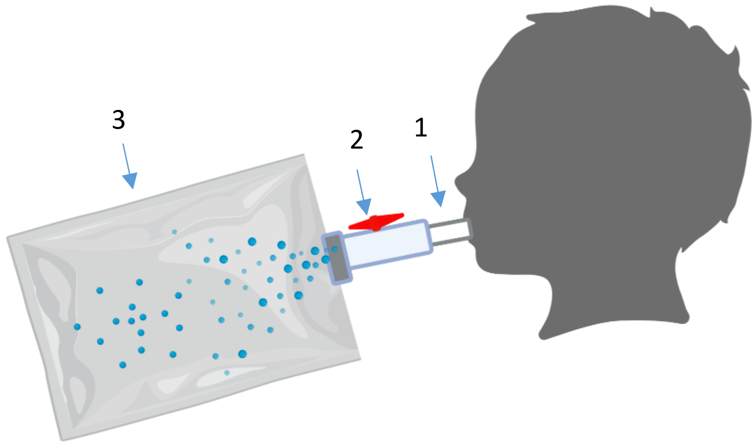


**Supplementary Figure 1. Breath collection system for children.** To collect breath, child places mouthpiece (1) between the lips and exhales completely. Volatiles are transferred from two-way valve (2) to SamplePro FlexFilm sample bag (3). Figure created with Biorender.com.

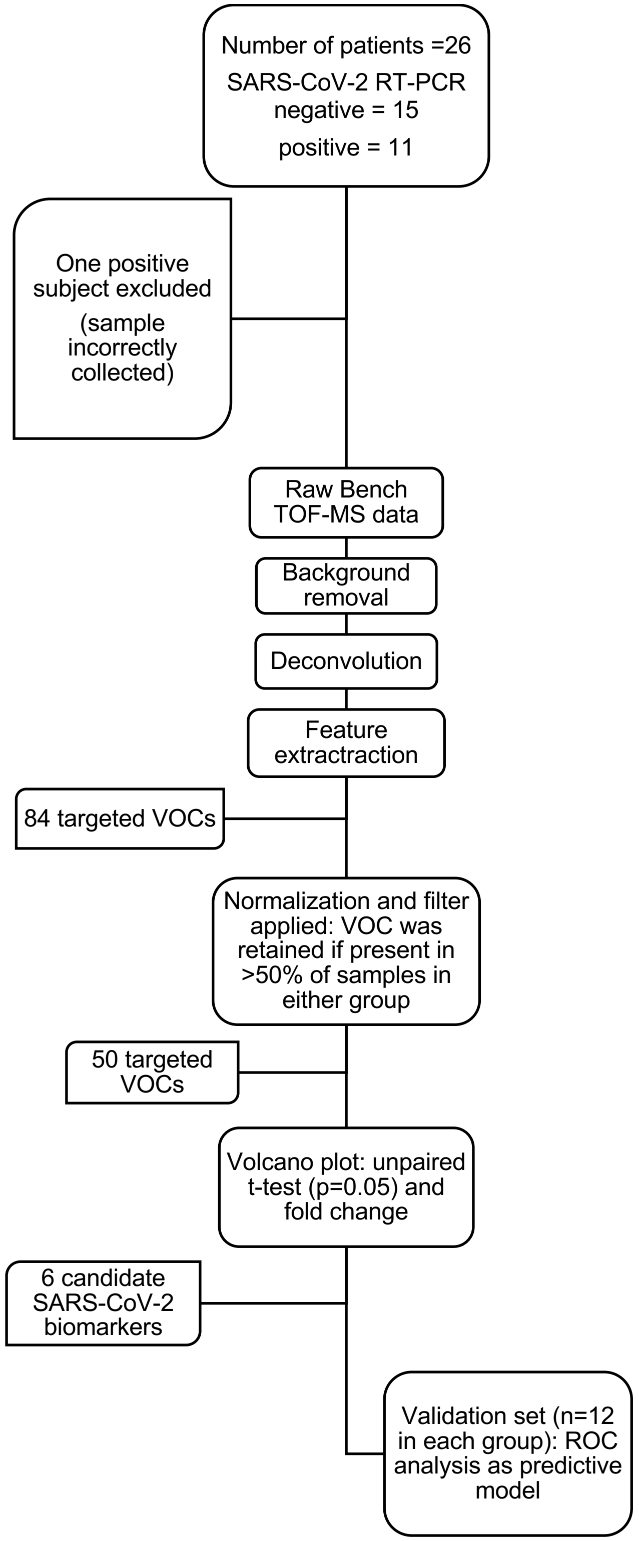


**Supplementary Figure 2: Workflow of data analysis and statistics used to create a final predictive model to discriminate SARS-CoV-2-infected from -uninfected subjects.**

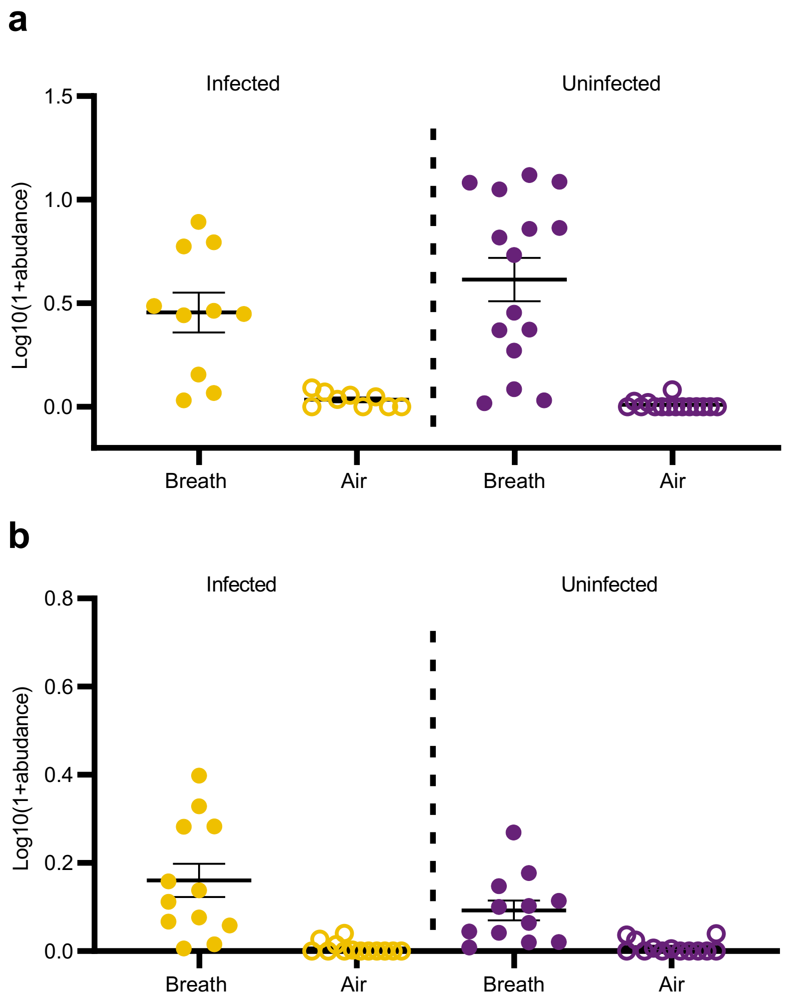


**Supplementary Figure 3. Isoprene is significantly more abundant in breath samples compared to room air [discovery cohort (a) and validation cohort (b)].** Higher levels of isoprene were found in breath compared room air in both SARS-CoV-2-infected and -uninfected breath samples. Median with SEM are shown. Note that the sensitivity of the instrument changed between the time of analysis of each cohort, as a result of system maintenance and recalibration.

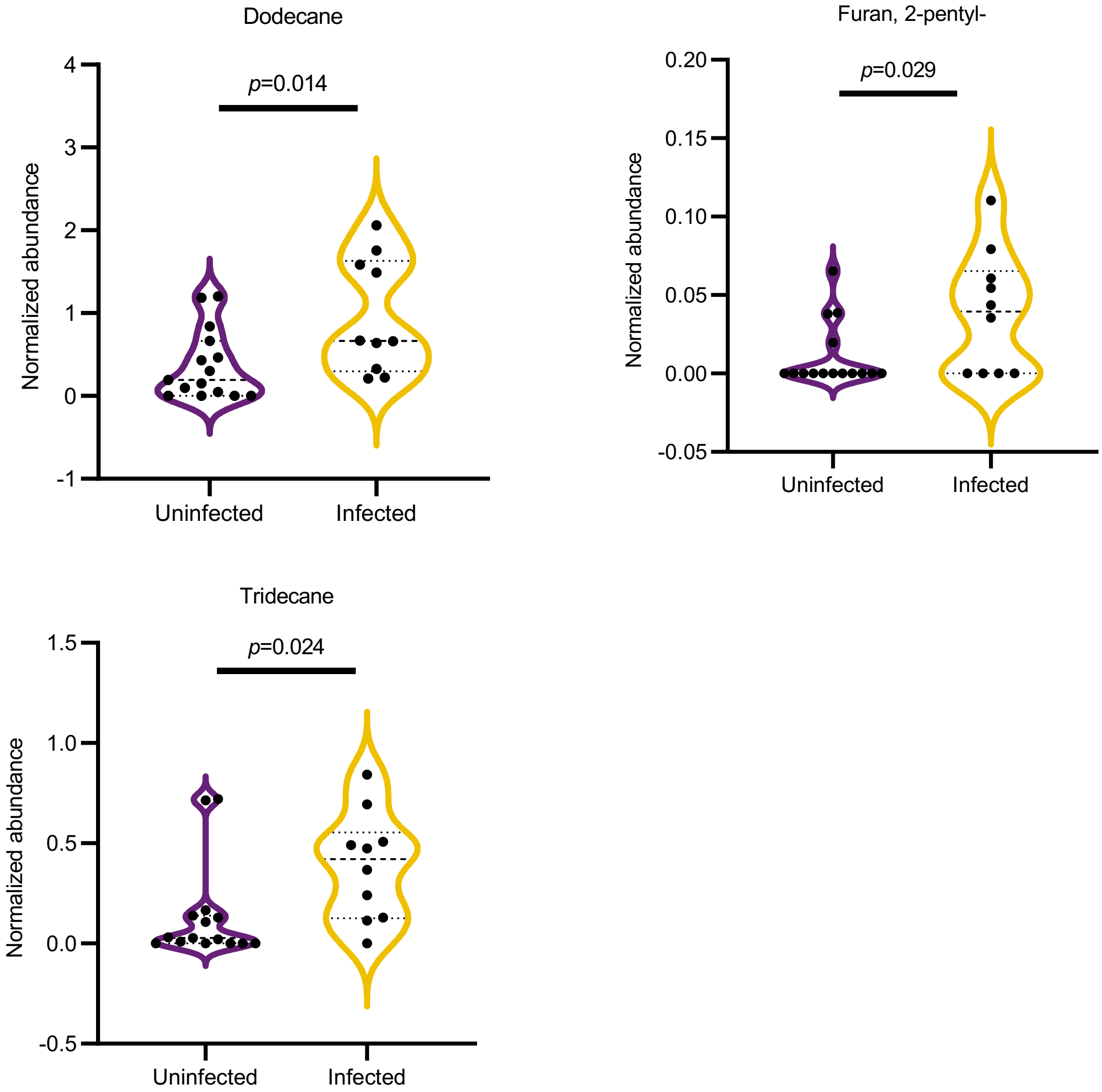


**Supplementary Figure 4. Breath abundance of candidate SARS-CoV-2 biomarkers in the breath of uninfected and infected children (discovery cohort).** Median and quartiles are shown. P-values from t-tests are shown for each comparison.

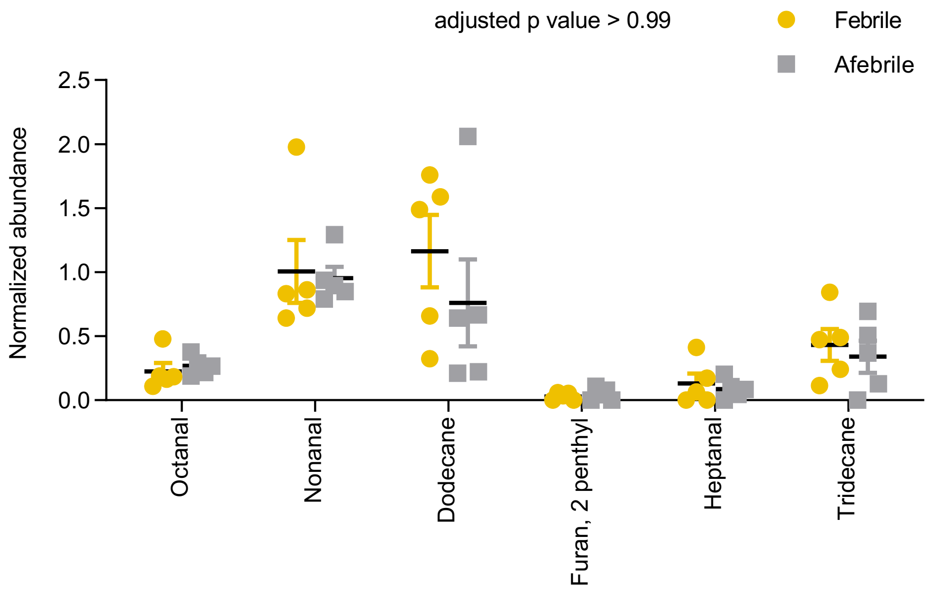


**Supplementary Figure 5. Breath abundance of candidate SARS-CoV-2-associated biomarkers are not significantly different between febrile and afebrile SARS-CoV-2 patients (discovery cohort).** Median with SEM are shown. Adjusted p value (t-test) for all comparisons >0.99.

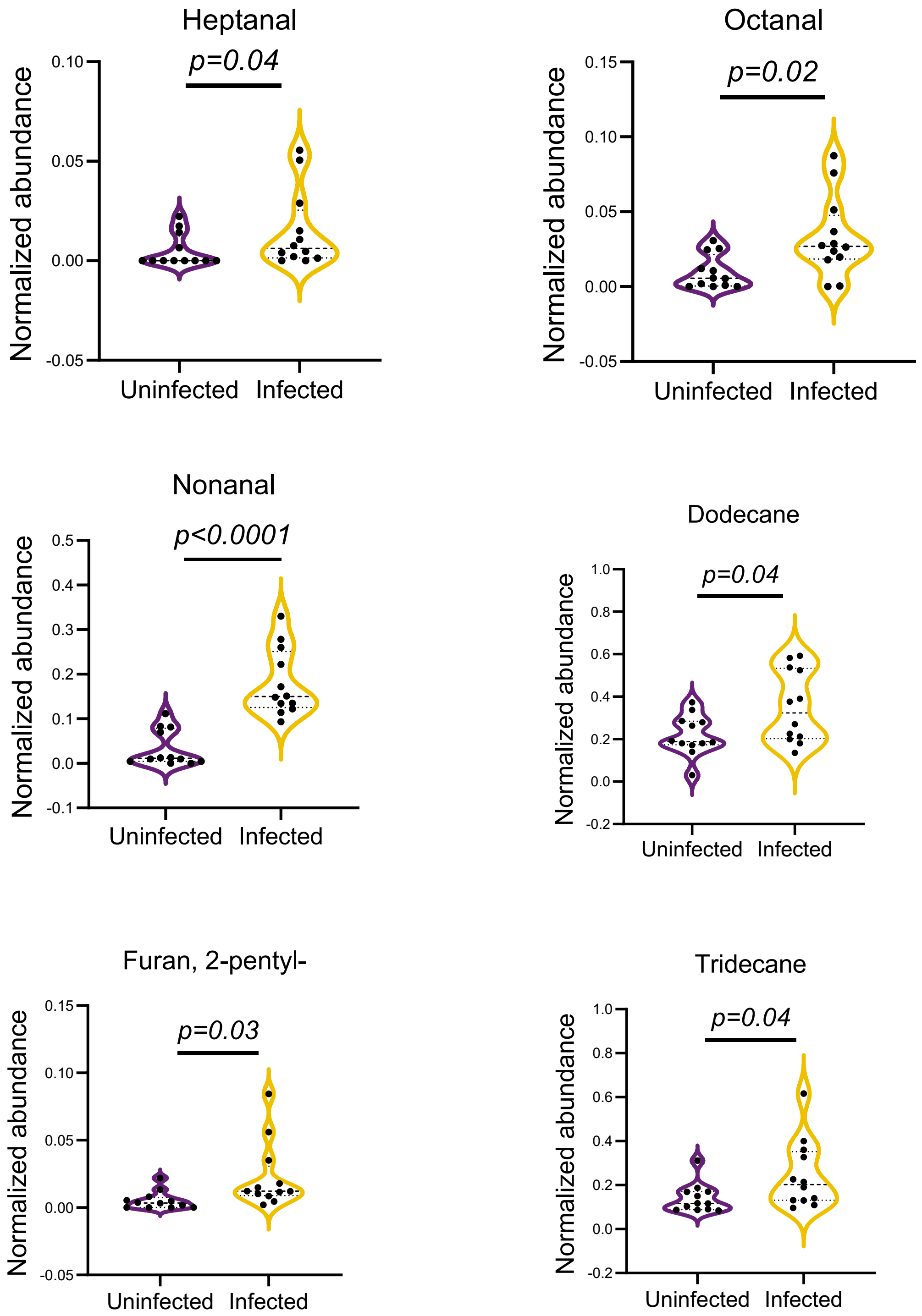


**Supplementary Figure 6. Breath abundance of candidate SARS-CoV-2 biomarkers in uninfected and infected children in the validation cohort.** Median and quartiles are shown. P-values (t-tests) are shown for each comparison.

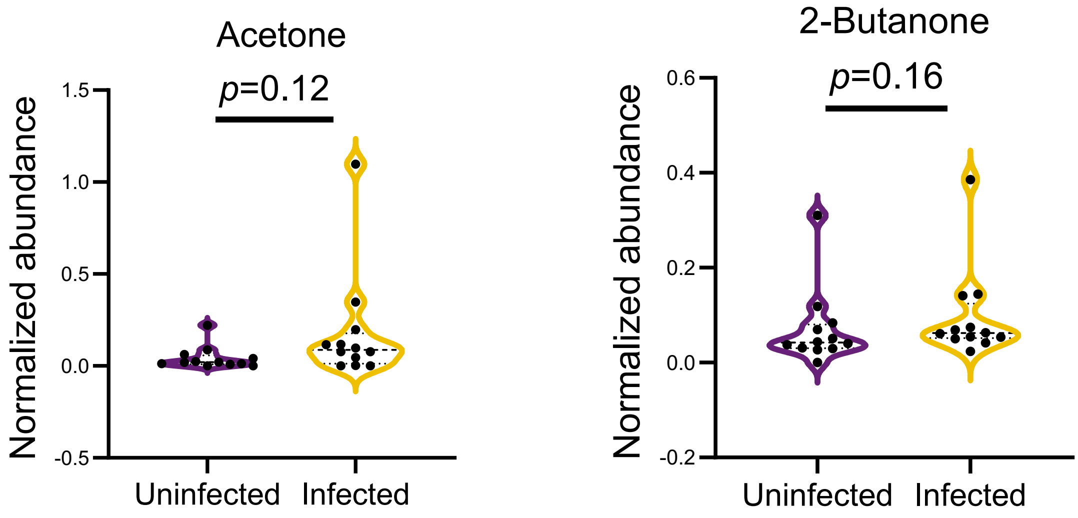


**Supplementary Figure 7. Ketones do not characterize the breath of pediatric SARS-CoV-2.** Acetone and 2-butanone previously reported in the breath of adults with and without COVID-19 10 were detected in our pediatric samples; however no significant differences were found between SARS-CoV-2 infected and uninfected pediatric samples in the validation cohort (similar to results in discovery cohort, Fig. 5). Median and quartiles are shown. P-values from t-tests are shown for each comparison.
